# Polygenic transcriptome risk scores enhance predictive accuracy in atopic dermatitis

**DOI:** 10.1101/2025.01.07.25320097

**Authors:** Charalabos Antonatos, Ashley Budu-Aggrey, Alexandros Pontikas, Adam Akritidis, Efstathia Pasmatzi, Aikaterini Tsiogka, Stamatis Gregoriou, Katerina Grafanaki, Lavinia Paternoster, Yiannis Vasilopoulos

## Abstract

**Background:** Incorporation of gene expression when estimating polygenic risk scores (PRS) in atopic dermatitis (AD) may provide additional insights in disease pathogenesis and enhance predictive accuracy.

**Objective:** In this study, we developed polygenic transcriptome risk scores (PTRSs) derived from AD-enriched tissues and evaluated their performance against traditional PRS models and a baseline risk model incorporating eosinophil and lymphocyte counts in the prediction of AD.

**Methods:** We conducted transcriptome-wide association studies (TWAS) using the PrediXcan framework to construct tissue-specific PTRSs. Risk score performance was assessed in 256888 Europeans (10,816 cases) and validated in an independent cohort of 64152 Europeans (2669 cases) from the UK Biobank.

**Results:** We observed a modest correlation between PRS and PTRS, exerting independent effects on AD risk. While PRS demonstrated superior predictive performance compared to single-tissue PTRSs, combining both models significantly enhanced prediction accuracy, yielding a c-statistic of 0.646 (95% confidence intervals: 0.634–0.656). Notably, tissue-specific PTRSs revealed stronger associations with baseline risk factors, where Eppstein-Bar virus (EBV)-transformed lymphocytes and unexposed skin PTRSs tissues reported positive associations with lymphocyte counts.

**Conclusion:** Our findings highlight the value of integrating transcriptome-based risk models to incorporating additional omics layer to refine risk prediction and enhance our understanding of genetic architecture of complex traits.

## Introduction

Atopic dermatitis (AD) is a widely prevalent skin disease that affects both infants and adults^1^. AD is characterized by recurrent eczematous lesions, intense itching, and a compromised skin barrier, which often leads to secondary infections. It is considered one of the earliest manifestations of the atopic march, a sequence of allergic diseases that typically begins with AD in childhood and may progress to asthma, allergic rhinitis, and other atopic disorders^1^. The genetic architecture of AD has been well-documented, with heritability estimates reaching as high as 80% in twin studies, indicating a substantial genetic contribution to the pathogenesis of the disease^2^. Over the past decade, genome-wide association studies (GWASs) have uncovered more than 100 genetic loci associated with AD, further highlighting its polygenic nature^3^.

Despite significant advancements in identifying risk loci for AD, much less attention has been given to identifying individuals at high risk. Polygenic risk scores (PRS) offer a promising approach for directly translating these findings into clinical practice^4^. PRS aggregates the effects of numerous genetic variants across the genome, each weighted by its effect size derived from large-scale GWASs, to estimate an individual’s genetic predisposition to a particular trait. PRSs have been widely applied for stratifying individuals based on their genomic profile, thus aiding the clinical practice for preventive measures4. Studies evaluating the performance of PRS in AD have shown, thus far, that PRS alone can adequately stratify individuals yielding high predictive values^5,6^.

Nonetheless, little effort has been made to incorporate additional layers of genetic information in constructing risk scores. A growing body of evidence suggests that many of the genetic variants associated with complex traits, such as AD, reside in non-coding regions of the genome, modulating gene regulatory processes that can influence disease susceptibility. An example of such regulatory interactions refers to *cis*-quantitative trait loci (*cis*-eQTLs), affecting gene expression variation^7^. Recent approaches have integrated genetically regulated gene expression (GReX) to risk score development through polygenic transcriptome risk scores (PTRS)^8^. PTRS leverage the cumulative effect of genes (here, at the expression level) to construct risk predictors. PTRS are based on the premise that gene expression changes driven by genetic variation are relatively stable across different populations and may therefore be more generalizable across diverse ancestry groups. Compared to traditional PRS, PTRS have been shown to offer improved portability across different traits and diseases^8,9^, providing a more robust and interpretable model of genetic risk. Moreover, gene-based scores not only capture the genetic variation associated with disease but also offer insight in the underlying molecular mechanisms of traits by incorporating gene expression. Hence, PTRS can reveal important aspects of the genetic architecture of complex traits like AD.

In this work, we developed PTRSs to examine the association between GReX and AD in UK Biobank (UKB) European participants. We compared the predictive performance of PTRSs to traditional PRS frameworks and a baseline risk score consisting of eosinophil and lymphocyte counts. We further constructed a combined risk score model that integrates both PRS and PTRS, assessing the predictive accuracy compared to single-risk scores. Finally, we explored the independent association between PTRS and clinical risk factors for AD, aiming to establish whether PTRS can further refine the prediction of AD risk beyond genetic risk alone. An overview of the study design is presented in Fig. 1.

**Fig. 1.**
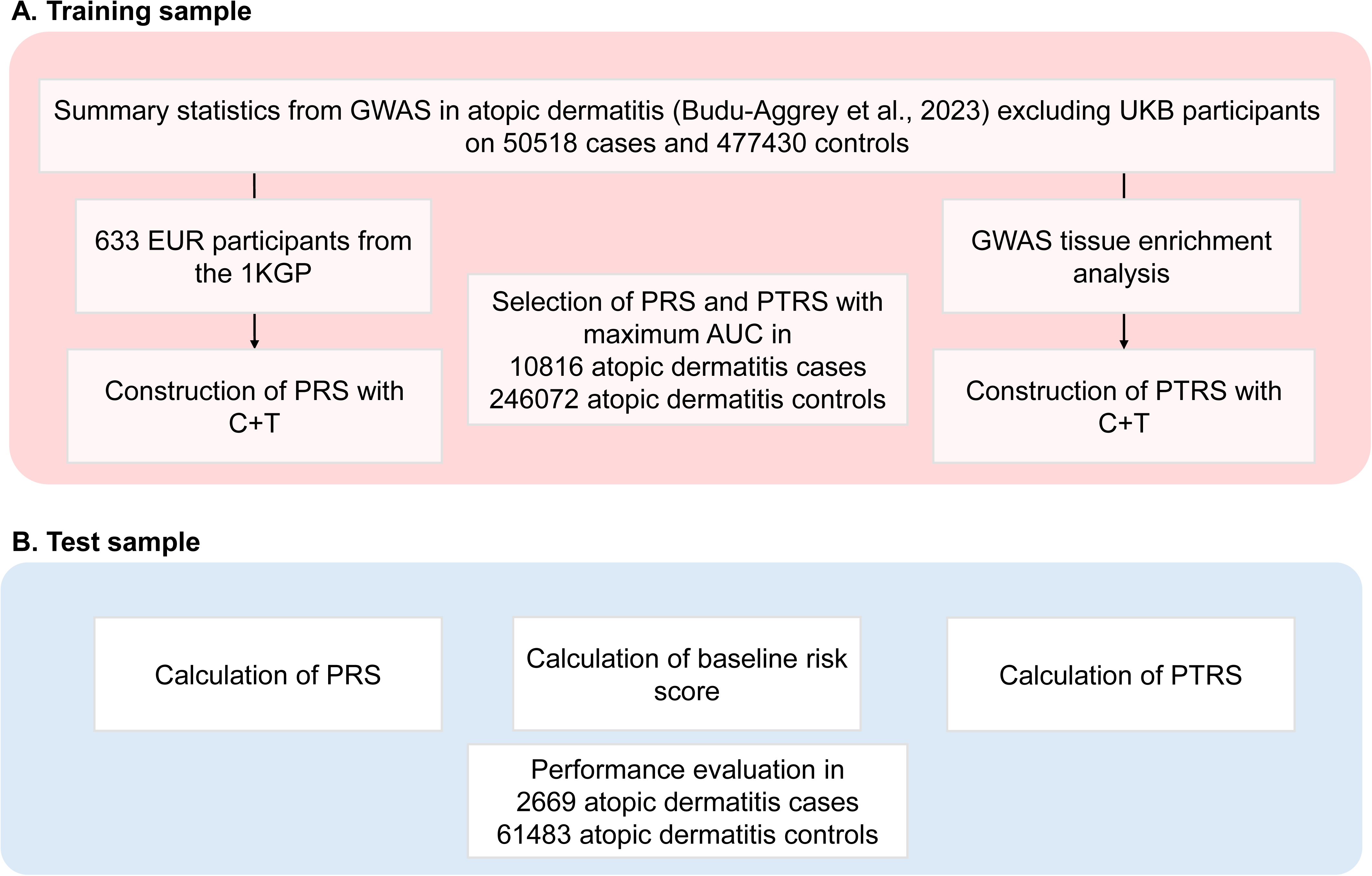
Study design. We selected the optimal parameters for each risk score in the training dataset (A) and evaluated the predictive accuracy in the training dataset (B). GWAS, genome-wide association study; UKB, UK Biobank; EUR, European; 1KGP, 1000 Genomes Project; PRS, Polygenic risk score; C+T, clumping and thresholding; PTRS, polygenic transcriptome risk score.

## Materials and Methods

### Data sources

All analyses were conducted on the GRCh38/hg38 human genome version. External GWAS data for AD were derived from 527948 European participants (50158 cases, 477430 controls) excluding UKB participants3. The meta-analysis was performed using GWAMA for 12142641 variants in a fixed effect model^10^. Analyses were subsequently restricted to common (minor allele frequency (MAF)>0.01), biallelic variants. Tissue-based expression models based on GTEx V8 data were retrieved from the PrediXcan database^11^. Linkage disequilibrium (LD) computations relied on an external reference panel of 633 European unrelated samples from the 1000 Genomes project (1KGP) reference panel^12^.

### Study participants

We used data from the UKB, a large-scale biomedical database containing genetic, lifestyle and health data from approximately half a million UK participants^13^. Genotyping of the participants was performed using the UKB Axiom Affymetrix array^13^. Genotypic data were lifted over from GRCh37 to GRCh38 using GATK Picard Liftover tool^14^ and consequently imputed from the Genomics England (GEL) 100,000 Genomes project with high-coverage sequence data^15^. The resulting GEL reference panel consisted of more than 300 million autosomal variants. Details regarding imputations and quality metrics are described elsewhere^15^.

Only European participants were included, with one random participant selected from each pair of at least third-degree relatives (kinship coefficient > 0.0884). AD cases were defined as individuals who self-reported “eczema/dermatitis” in a verbal interview during their initial visit at the assessment center (Data field ID: 20002). The rest of eligible participants were used as controls. Individuals listed as controls were excluded if they had previously self-reported that had hay fever, allergic rhinitis or eczema (Data field ID: 6152). We randomly split the eligible participants into a 80% training set to evaluate the performance of PRS and PTRS, and 20% testing set to apply the optimal risk score maintaining the same ratio for age, sex and case/control status.

### Tissue enrichment of AD GWAS

To select eligible tissues for PTRS computations, we performed a gene property analysis in the functional mapping and annotation of GWASs (FUMA) platform v1.5.2^16^ using MAGMA v1.10^17^. Briefly, MAGMA conducts a gene-based association test producing a one-sided P-value. At next, gene-based P-values are transformed to Z-scores and are associated with expression values from different tissues. We selected 49 pre-computed GTEx v8 tissue expression estimations with available *cis*-eQTLs^7^ and conducted a one-sided test to prioritize AD-relevant tissues based on gene-level results. A Bonferroni-corrected P-value threshold of 0.05/49 was adopted to declare significant results.

### Derivation of polygenic transcriptome risk score

We selected significantly enriched tissues from the tissue enrichment analysis of AD GWAS present in the PrediXcan database. Summary-based TWASs were conducted using the S-PrediXcan approach to estimate the effect size of each gene^18^. We used multivariate adaptive shrinkage models based on fine-mapped variables from deterministic approximation of posteriors (MASHR-M)^19^ for enriched GTEx tissues as per authors’ recommendations^20^. The above models are mapped to the GRCh38/hg38 human genome version and are restricted to a small number of available *cis*-eQTLs, resulting in decreased intersections between variants available in the prediction model and GWAS and thus reduced performance. Therefore, we (i) harmonized the base GWAS variants through liftOver to the GRCh38/hg38 human genome version, and (ii) imputed the base GWAS variants in a region-wide approach to increase the available number of intersected SNPs. Both steps were run according to the S-PrediXcan pipeline^21^.

Next, we calculated tissue-specific PTRS through the PrediXcan framework^11^. In the original implementation of PTRS, the estimated effect of a gene was calculated using the GReX as feature through elastic net models^9^. Here, we constructed PTRSs using a summary statistics-based method, where per-gene effects were derived from S-PrediXcan. In particular, for an *i^th^* individual, we compute the PTRS as:

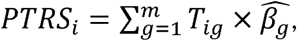

where *T_ig_* is the GReX of a gene *g* in the *i^th^* individual estimated through the PrediXcan framework, and 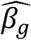 is the estimated effect of a gene g estimated from the S-PrediXcan framework. PTRSs were calculated for sequential P-value thresholds including a different number of genes in each case, referring to P-value=1, P-value≤0.1, 5×10^-2^, 5×10^-3^, 5×10^-4^, 5×10^-5^, 5×10^-6^, 5×10^-7^ and 1×10^-7^. Each PTRS was standardized prior to evaluation in both training and test datasets and adjusted for age, sex, and the first 10 genetic principal components.

### Polygenic risk scores for AD

To compare the proposed PTRS compared to a traditional PRS approach, we calculated PRS using clumping and thresholding (C+T) in the UKB. The C+T approach involves a clumping algorithm to yield an independent number of SNPs selecting those mostly associated with the phenotype. Variants were clumped using an external 1KGP European LD reference panel^12^. Next, we constructed PRSs using the PRSice-2 v2.3.5 software^22^. All derived PRSs were adjusted for age, sex, 10 first genetic principal components and standardized prior to evaluation in training and test datasets.

### Statistical analyses

We first formed a baseline risk model for AD based on eosinophil count (Data-field ID: 30150), lymphocyte count (Data-field ID: 30120), age and sex. Blood cell count phenotypes were rank-based inverse normal transformed^23^. Missing values were imputed using multivariate imputation by chained equations with random forest (MICE)^24^. We calculated the odds ratio (OR) and 95% confidence intervals for eosinophil and lymphocyte counts using logistic regression.

Evaluation of standardized PRS and PTRSs was performed using the maximal AUC approach. Predictive ability of each method was assessed with the receiver operator characteristics (ROC) curve by computing Harrel’s c-statistic and 95% confidence intervals (95% CIs) using Delong’s method via 10000 stratified bootstraps. The c-statistic estimates the likelihood that a randomly selected case has a higher risk score than a randomly selected control. C-statistic values range from 0.5 (random classification) to 1 (perfect classification). Pairwise comparisons between ROC curves were conducted using Delong’s method^25^. C-statistics and Δc-statistics with corresponding 95% confidence intervals (95% CIs) were performed using the pROC R package^26^. Risk scores were adjusted for age, sex, and the first 10 genetic principal components. We chose the best performing PRS and PTRS model in the training dataset and applied it to the test dataset.

A combined risk score for standardized PRS and PTRS was calculated with a weighted sum. Pearson correlation coefficients between the best-performing standardized PRS and PTRSs as well as their interactions were estimated through logistic regressions in the train dataset. Interaction estimates were adjusted for age, sex and 10 first genetic principal components. We prioritized standardized PTRSs that showed non-significant interactions with standardized PRS in the train dataset and evaluated the predictive accuracy in the test dataset. We evaluated the overall performance of risk scores using (i) a baseline risk model consisting of age, sex and blood cell counts, (ii) a risk score approach for each standardized PRS and PTRS alone, (iii) a combination of the baseline risk model and standardized PRS/PTRS, and (iv) a combination of baseline risk model, standardized PRS and PTRS in a tissue-specific manner.

We finally assessed the interaction of standardized PRS and PTRSs in association with blood cell counts using linear regression in the test dataset. Rank-based inverse normal transformed blood cell counts, including eosinophil and lymphocyte counts were included as outcome variables. Interactions between lymphocyte counts and standardized PTRSs in AD risk were assessed by adding an interaction term in the regression analysis. All estimates were adjusted for age, sex, and first 10 genetic principal components to account for population structure. Derived P-values for interaction analyses were adjusted for Bonferroni correction.

## Results

### Participant characteristics

Our study included 321040 unrelated participants of European ancestry from the UKB. Based on self-reporting data and matched case/control, age and sex splitting, training data comprised 10816 AD cases and 246072 controls, while test data included 2669 AD cases and 61483 controls. Demographic and clinical characteristics are presented in Table 1.

**Table 1.** Descriptive statistics of study participants.

|  | Training dataset |  | Test dataset |  |
| --- | --- | --- | --- | --- |
|  | Cases | Controls | Cases | Controls |
| Age (mean ± SD) | 55.276<br>(8.158) | 57.405 (7.915) | 55.256<br>(8.140) | 57.406<br>(7.913) |
| Sex (Male/Female) | 4943/5873 | 116095/129977 | 1221/1448 | 29009/32474 |
| Eosinophil counts (mean ± SD) | 0.204<br>(0.181) | 0.167 (0.129) | 0.203<br>(1.629) | 0.166<br>(0.128) |
| Lymphocyte counts (mean ± SD) | 1.894<br>(1.546) | 1.961 (1.781) | 1.891<br>(0.759) | 1.958<br>(1.259) |
| Units of measurement for blood cell counts are $10^9$ cells/Litre. Abbreviations: SD, standard deviation. | | | | |

### Transcriptome-wide analyses for AD

Enrichment analysis across 49 GTEx tissues with available *cis*-eQTL data revealed significant associations in 7 tissues, namely whole blood (P-value=2.37×10^-10^), spleen (P-value=9.78×10^-9^), Epstein-Barr virus (EBV) transformed lymphocytes (P=value=7.58×10^-8^), small intestine (P-value=8.14×10^-6^), not sun (P-value=2.32×10^-4^) and sun (P-value=4.51×10^-4^) exposed skin and lung (P-value=4.54×10^-4^) tissues (Fig. 2a). These findings align with prior tissue enrichment analyses in AD GWASs^3^, highlighting the multi-tissue etiological mechanisms underlying AD pathogenesis and possible links to the atopic march. By applying the S-PrediXcan framework in each of the 7 tissues, we identified 175 genes in total (72 unique) associated with AD risk at a Bonferroni corrected P-value threshold of 5.79×10^-7^ (Fig. 2b). The total number of S-PrediXcan results is provided at Tables S1-S7.

**Fig. 2.**
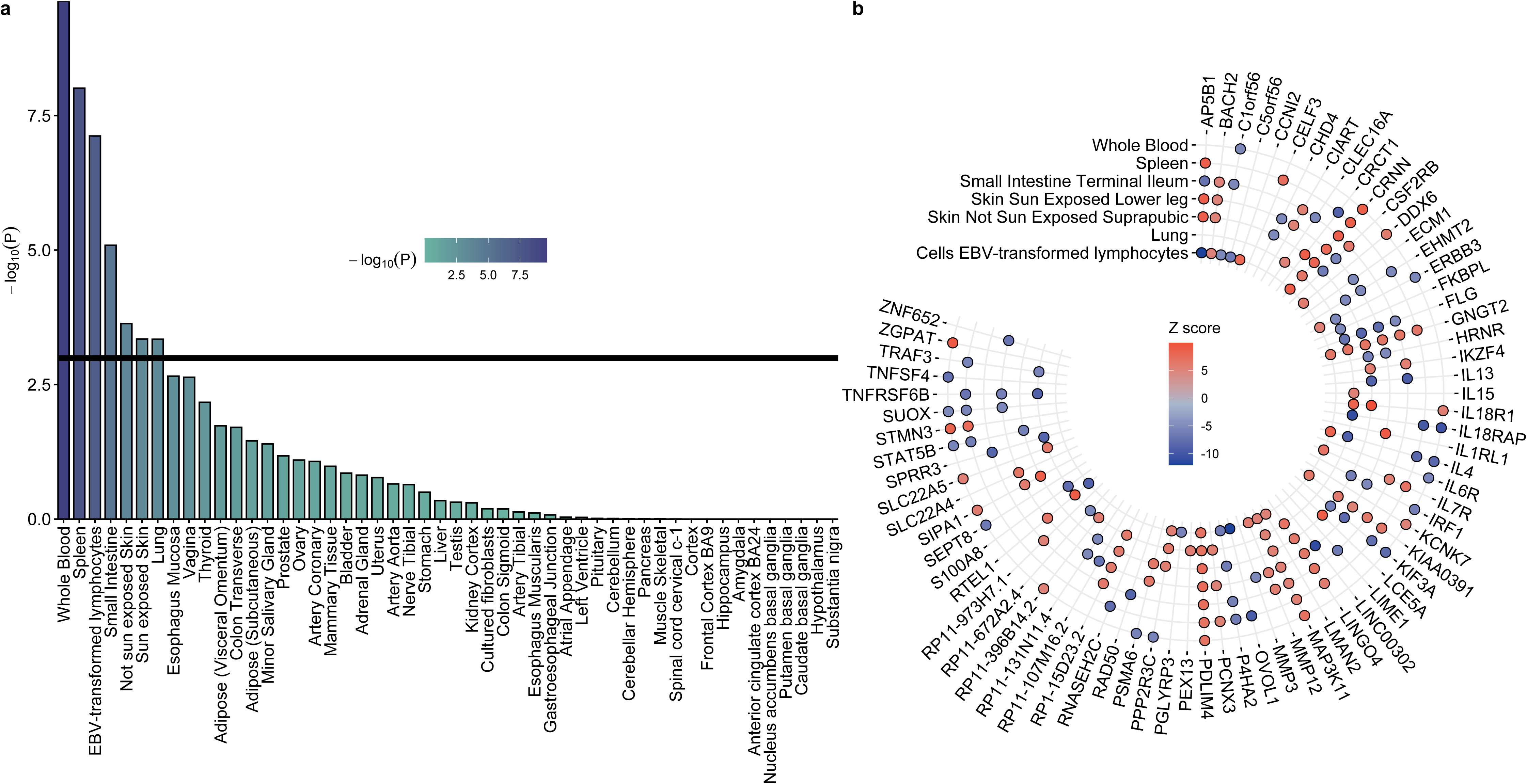
Tissue enrichment analysis on atopic dermatitis and significant genes in each tissue. (a) Significantly enriched tissues in the atopic dermatitis GWAS. (b) Transciptome-wide association analysis results for each of the 7 statistically significant tissues (Tables S1-S7).

### Selection of the best performing risk score and assessment of a baseline risk score

The optimal PRS and PTRS models were selected using 256888 unrelated participants of European ancestry (10816 cases; Table 1). For standardized PRS, the best performing model was estimated through PRSice2 v2.3.5 at a P-value threshold of 5.005×10^-5^ including 390 SNPs in total (Fig. S1, Table S8). On the contrary, we chose the PTRS model in each GWAS-enriched GTEx tissue through the maximal AUC approach. No specific pattern of gene number arose in the training data, suggesting a tissue-specific effect of standardized PTRS in AD risk (Table S9). For example, the maximum number of genes was reported at whole blood at a P-value<0.005 threshold (n=255), while small intestine incorporated the lowest number of genes at a P-value<5×10^-7^ threshold (n=23; Table S9). Among the PTRS models, the highest discriminative ability was observed in not sun exposed skin (c-statistics, 95% CI: 0.599, 0.594-0.605) using 106 genes.

We next evaluated the magnitude of strength of association of clinical risk factors for AD. Eosinophil counts were positively associated with AD risk (log(OR), 95% CI: 0.268, 0.249-0.288; P-value<2×10^-16^), while lymphocyte counts were negatively associated with AD risk (log(OR), 95% CI: -0.110, -0.129-0.090; P-value<2×10^-16^). These results are in line with previous reports confirming the established association of eosinophils in AD risk and severity^27^, while patients with AD have in general lower lymphocyte counts compared to healthy controls^28^. Their discriminative abilities were 0.601 (95% CI: 0.596–0.607) and 0.581 (95% CI: 0.575– 0.586), respectively, combining for a baseline risk model with a c-statistic of 0.611 (95% CI: 0.606–0.617).

### Predictive accuracy of PRS and PTRS

In the test dataset comprised of 2669 cases and 61483 controls, we examined the performance of standardized PRS and PTRS scores in predicting AD risk (Table 1). The baseline model comprised of age, sex, eosinophil and lymphocyte counts yielded a c-statistic of 0.616 (0.605-0.628; Fig. 3). The PRS model alone demonstrated the highest overall accuracy (c-statistic, 95% CI: 0.619, 0.608-0.630; Fig. 3). Among standardized PTRS models, sun exposed skin showed the strongest predictive ability (c-statistic, 95% CI: 0.604, 0.593-0.615; Table S10). The standardized PRS significantly outperformed the best standardized PTRS model (Δc-statistic, 95% CI: 0.015, 0.007-0.02; P-value=6.61×10^-5^), however showing comparable performance to the baseline risk score (Δc-statistic, 95% CI: 0.002, -0.008-0.014; P-value=0.645).

**Fig. 3.**
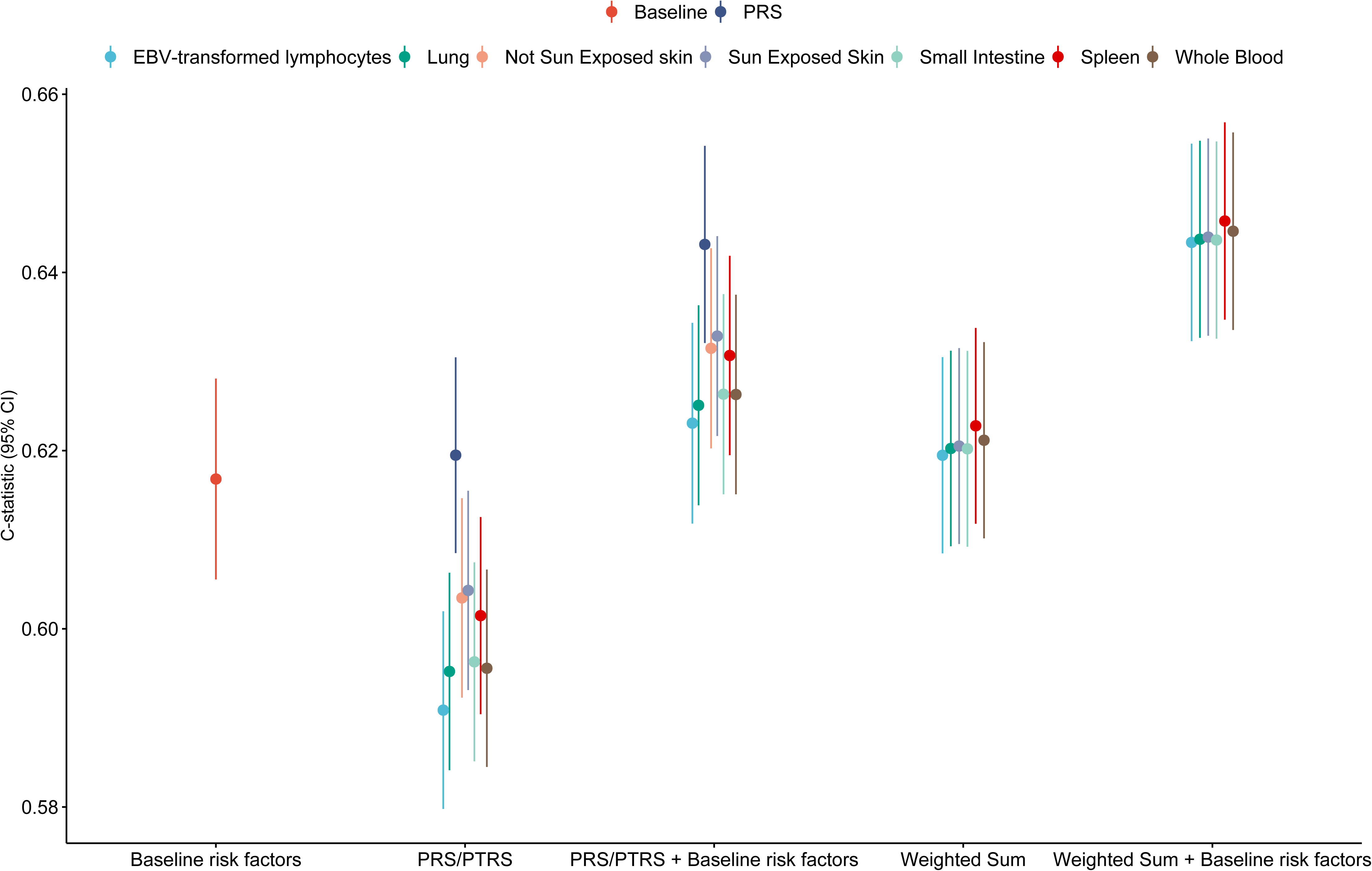
Predictive accuracy of polygenic risk scores and polygenic transcriptome risk scores in the test dataset (Table S10). The y-axis represents the estimated c-statistic with accompanying 95% confidence intervals. Color indicates each separate method used to calculate the c-statistic. CI, confidence intervals; PRS, polygenic risk score; PTRS, polygenic transcriptome risk score.

A similar pattern of association derived when adding the baseline risk model in standardized PRS/PTRS risk scores. Despite significant improvements of the standardized PTRS models compared to standardized PTRSs alone (Fig. 3), the standardized PRS remained the most superior model when compared to the best performing PTRS model (Δc-statistic, 95% CI: 0.010, 00.004-0.015; P-value=3.46×10^-4^).

### Combined PRS and PTRS

Both risk scores were further evaluated for their combined predictive utility. PRS and each tissue-specific PTRS model reported significant correlations (Figs. S2-S8), nevertheless reporting non-significant interactions (Table S11) excluding standardized PTRS for not sun exposed skin (P-value=1.08×10^-4^; Table S11). Hence, excluding not sun exposed skin, standardized PTRS provides an additional layer of genetic risk information that may be useful for stratification. We calculated weights for each PRS/PTRS comparison through logistic regression in the training dataset, excluding not sun exposed skin, and evaluated the predictive accuracy of the PRS and PTRS in the test dataset through weighted sum.

As expected, the weighted sum of standardized PRS and PTRS outperformed standardized PRS and PTRS alone (Fig. 3). The best performing model in weighted sum was reported in spleen (c-statistic, 95% CI: 0.622, 0.611-0.633) with significant differences compared to standardized PRS alone (Δc-statistic, 95% CI: 0.003, 0.001-0.005; P-value=0.001) and standardized PTRS in sun exposed skin (Δc-statistic, 95% CI: 0.018, 0.011-0.025; P-value=3.19×10^-7^). When incorporating baseline risk factors, the c-statistic reached a value of 0.646 (95% CI: 0.634-0.656), surpassing the best performing model in the single-risk analysis (Fig. 3). For instance, the difference in predictive accuracy between weighted sum and clinical risk factors was significant compared to standardized PRS and clinical risk factors (Δc-statistic, 95% CI: 0.002, 0.001-0.004; P-value=5.59×10^-4^). A complete description of c-statistics and corresponding 95% CIs is provided at Table S11.

### Association with disease severity

Given the lack of clinical metrics for AD in the UKB cohort, we hypothesized that previously associated clinical risk factors for AD could be used as proxies. Increased eosinophil counts have been long associated with AD onset and severity, while patients with AD report lymphopenia. Hence, we assessed the association of standardized PRS and each tissue-specific PTRS scores in rank-based inverse normal transformed eosinophil and lymphocyte counts. The interaction was computed by adding an interaction term in the same prediction model for evaluation.

Both standardized PRS and PTRS scores showed significant associations with eosinophil counts, with independent contributions to the distribution of the latter except for sun exposed skin standardized PTRS (Fig. 4a; Table S12). However, the standardized PTRSs for EBV-transformed lymphocytes (log(OR), 95% CI: 0.031, 0.023-0.040), not sun exposed skin (log(OR), 95% CI: 0.036, 0.027-0.045) and small intestine (log(OR), 95% CI: 0.043, 0.035-0.052) tissues reported an increased strength of association with eosinophil counts compared to standardized PRS (Fig. 4a; Table S12). Contrastingly, a more distinct association pattern was observed in lymphocyte counts, where the standardized PTRS derived from EBV-transformed lymphocytes (log(OR), 95% CI: 0.015, 0.007-0.023) and not sun exposed skin (log(OR), 95% CI: 0.017, 0.008-0.025) tissues were the only associations reaching significance threshold (Fig. 4b). The contradictory association patterns between lymphocyte counts and PTRSs (Fig. 4b) compared to their associations with AD risk prompted us to investigate their interactions in disease risk. Notably, standardized PTRSs in EBV-transformed lymphocytes and not sun exposed skin showed independent associations with AD risk compared to lymphocytes (Table S13), highlighting distinct interactions between lymphocyte-related mechanisms and genetic risk.

**Fig. 4.**
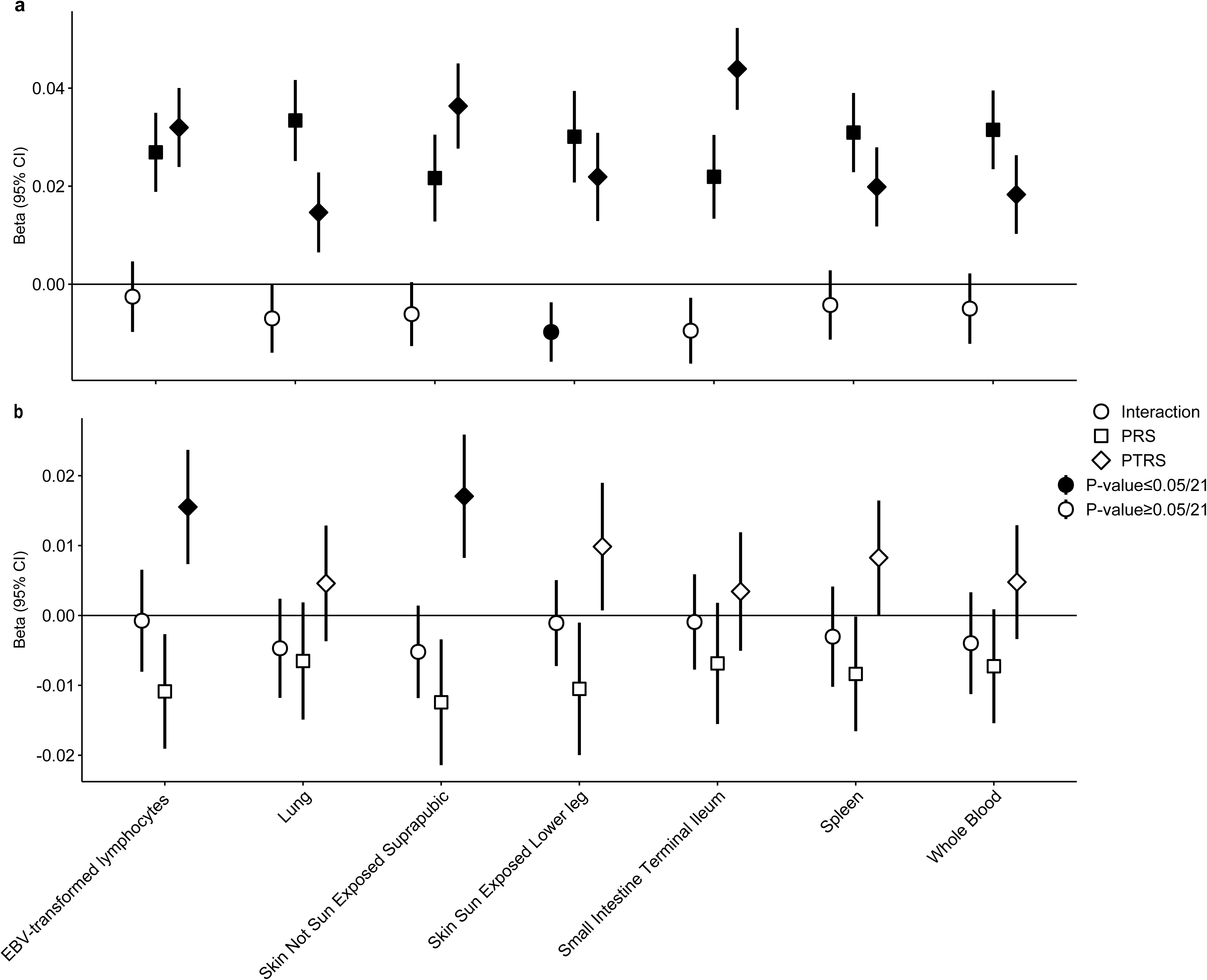
Interaction analysis of polygenic risk score and polygenic transcriptome risk score with baseline predictors. Each shape represents an estimate. Nodes with black fill represent P-values passing the Bonferroni-corrected significance threshold (P-value≤0.05/21). (a) Interaction analysis of polygenic risk score and polygenic transcriptome risk score with eosinophil counts (Table S11). (b) Interaction analysis of polygenic risk score and polygenic transcriptome risk score with lymphocyte counts (Table S12).

## Discussion

Here, we developed transcriptome-based polygenic risk scores to predict AD risk. By adding an additional layer of tissue-specific biological information to conventional genetic models, we aimed to disentangle gene- and tissue-specific contributions to genetic risk. We hypothesized that leveraging predicted gene expression variability in relevant tissues would enhance our understanding of AD pathogenesis and subsequently prediction risk. To assess this, we compared the predictive performance of PTRSs to traditional PRS frameworks and assessed the magnitude of strength of association with a baseline clinical risk score.

To refine our framework, we performed tissue enrichment analyses on the base GWAS data, identifying 7 significant tissues (Fig. 2a). While AD associations were evident in tissues such as skin and immune-related backgrounds (e.g., whole blood, spleen, and EBV-transformed lymphocytes), the observed enrichment in lung and small intestine was particularly notable. The enrichment in the above tissue stem from the generalized atopic background of AD, given the shared inflammatory pathways driving the atopic march, and the critical role these tissues hold in AD pathogenesis. For instance, studies have shown that infants with AD show compromised lung functionality independently of disease severity and food sensitivity^29^. Moreover, transcutaneous sensitization has been implicated in modulating food allergy risk^30^, further linking these tissues to the broader atopic phenotype.

In line with previous PRS applications in AD^5,6^, we observed that PRS alone outperformed baseline risk factors associated with AD in terms of maximal AUC, while inclusion of the latter enhanced the predictive ability. The underperformance of PTRS compared to both baseline risk models and PRS is likely due to the limited, fine-mapped *cis*-eQTL variants incorporated during PTRS weight construction^20^ compared to the reliance of PRS on genome-wide variants (Fig. 1). Comparisons between tissue-specific PTRSs must also be interpreted with caution given the distinct *cis*-eQTLs and biological contexts of each tissue. Previous reports have already shown that PTRSs outperform PRS in cross-ethnic portability, with improved association scores in chronic pulmonary obstructive disease^9^ and quantitative traits^8^. This advantage may arise from shared disease biology across ancestry groups, and the incorporation of cross-ancestry prediction models during PTRS construction^7^. Here, PTRSs demonstrated significant associations with larger, independent effect estimates compared to PRS in baseline risk factors (Fig. 4), unveiling a tissue-specific interaction of gene expression regulation and AD risk. We assume that PTRS weights for AD risk capture a broader inflammatory profile and immune activation in a tissue-specific manner. For instance, the independent association of small intestine PTRS in eosinophil counts has been previously suggested by functional studies, where increased eosinophils in the small intestine after allergic sensitization resulted in AD skin inflammation^32^. Similarly, EBV-transformed lymphocyte and not sun exposed skin PTRSs were independently, positively associated with lymphocyte counts (Fig. 4b). While this might appear contradictory, given that reduced lymphocyte counts were associated with AD, the results suggest that high PTRSs reflect a primed immune state specific to the studied tissues. Furthermore, the independent effects of PTRSs on AD risk (Table S13) support the notion that these scores capture distinct biological pathways related to immune activation, with compensatory effects that do not directly exacerbate disease risk. Thus, PTRSs may reflect tissue-specific biological mechanisms in disease risk. Functional studies could explore whether gene sets included in each PTRS model directly contribute to AD risk, however this falls outside the scope of this manuscript.

Our study has caveats. First, reliance on self-reported AD data in UKB introduces potential misclassification bias compared to clinical-grade diagnostic information. Secondly, we included only participants of European ancestry, thus limiting the generalizability of our results. Despite the established cross-ethnic portability of PTRSs, their association with clinical risk scores is yet to be uncovered. We suspect that this portability may not hold when assessing clinical risk scores in AD, due to ancestry-specific molecular mechanisms^32^. Third, stratification of risk scores per serum IgE levels, reflecting extrinsic (high, allergen-specific IgE levels) and intrinsic (normal IgE levels) AD endotypes was not feasible in this study. Fourth, our study included limited risk factors relevant for AD, focusing on well-established blood cell counts (Fig. 4). Using AD-enriched cohorts, as in prior PRS studies^5^ and incorporation of additional risk factors for AD (e.g., parental atopic history)^33,34^ is indispensable to further assess the utility of PTRSs, a goal we aim to pursue in future research. Similarly, while our findings provide a framework for risk prediction, its clinical application would primarily target much younger individuals where AD onset often occurs. This age difference may introduce variability in the model’s predictive performance due to distinct risk factors and disease mechanisms in younger populations. Lastly, the presented framework can be expanded to integrate additional omics layers, especially for genes that exert their effects on AD risk via mechanisms beyond gene expression^35,36^.

In conclusion, we constructed transcriptome-based polygenic risk scores for AD and evaluated their performance in UKB. Although standard PRS frameworks showed superior performance compared to all models, we revealed that tissue-specific PTRS scores provide unique biological insights by capturing tissue-relevant regulatory mechanisms underlying AD risk. Notably, PTRS models derived from AD-related tissues, such as EBV-transformed lymphocytes and not sun exposed skin, highlighted distinct interactions with lymphocyte counts, thus suggesting stratification based on tissue-specific contributions. These findings advance our understanding on the genetic architecture of AD and related systemic manifestations and provide the framework for integration of additional omics data. Future studies could expand on these insights to characterize the underlying molecular mechanisms governing each tissue and refine personalized risk prediction.

## Data Availability

All data produced in the present study are available upon reasonable request to the authors.

## Acknowledgements

This research has been conducted using the UK Biobank Resource under Application Number 285697. This work uses data provided by patients and collected by the NHS as part of their care and support. The analysis presented in this paper makes use of GWAS summary statistics from the EAGLE eczema consortium^36^. Charalabos Antonatos was financially supported by the “Andreas Mentzelopoulos Foundation”.

## Author Contributions

CA and YV contributed to the conception and design of the study. CA, ABA, AA, LP, YV contributed to acquisition of data and CA, ABA, AA performed statistical analysis of data. YV supervised the project. All authors contributed to the drafting and critical revision of the manuscript.

