## Supplementary material for "Polygenic transcriptome risk scores enhance predictive accuracy in atopic dermatitis": Fig. S

**Supplementary Figures**

Fig. S1. High-resolution PRSicev2 plot reporting the predictive accuracy of PRS across various P-value thresholds in the training dataset.

Fig. S2. Pearson correlation estimates between standardized PRS and standardized PTRS in EBV transformed lymphocytes.

Fig. S3. Pearson correlation estimates between standardized PRS and standardized PTRS in lung.

Fig. S4. Pearson correlation estimates between standardized PRS and standardized PTRS in not sun exposed skin.

Fig. S5. Pearson correlation estimates between standardized PRS and standardized PTRS in sun exposed skin.

Fig. S6. Pearson correlation estimates between standardized PRS and standardized PTRS in small intestine.

Fig. S7. Pearson correlation estimates between standardized PRS and standardized PTRS in spleen.

Fig. S8. Pearson correlation estimates between standardized PRS and standardized PTRS in whole blood.

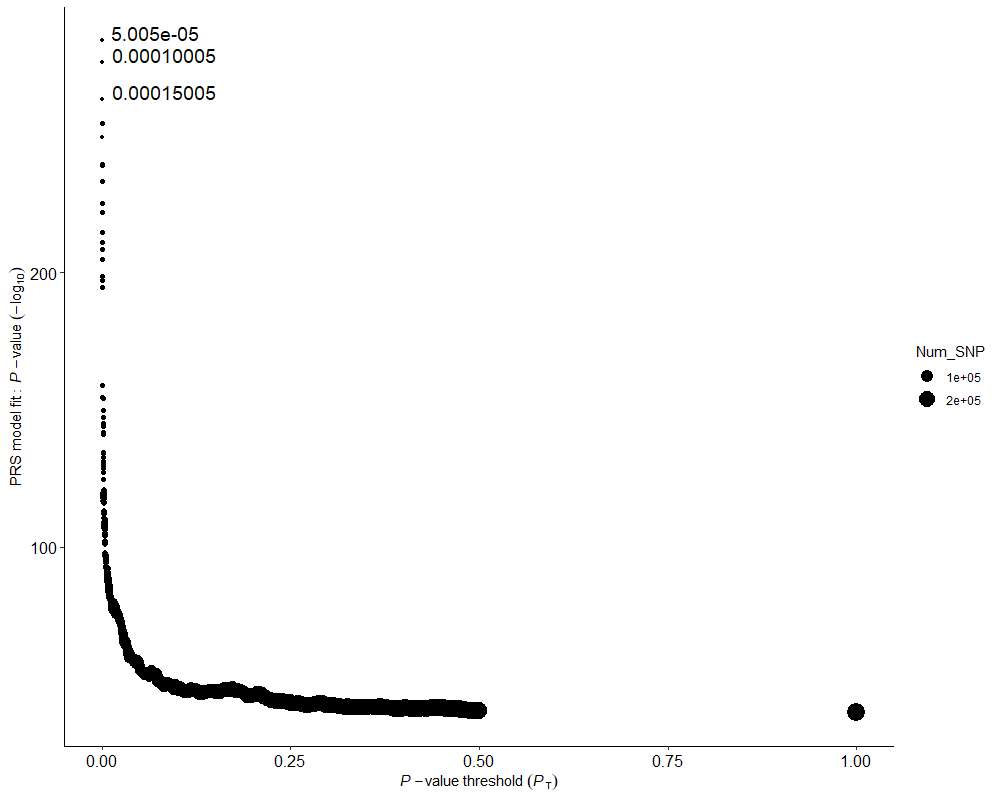
Fig. S1. High-resolution PRSicev2 plot reporting the predictive accuracy of PRS across various P-value thresholds in the training dataset.

Fig. S2. Pearson correlation estimates between standardized PRS and standardized PTRS in EBV transformed lymphocytes.

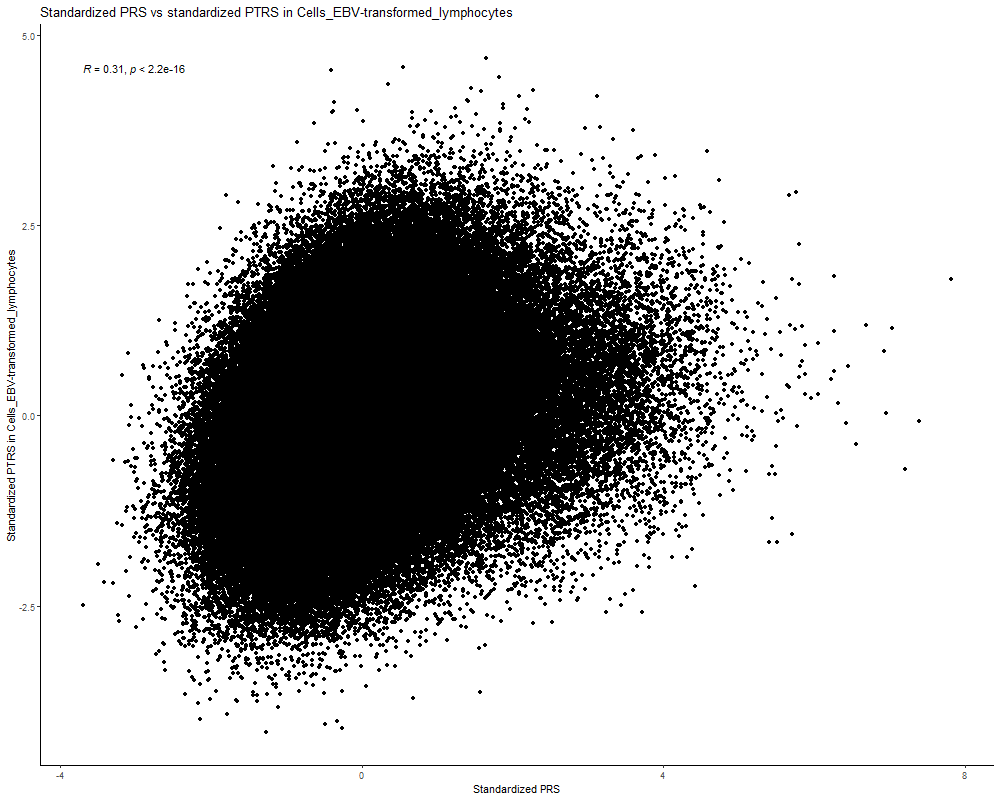

Fig. S3. Pearson correlation estimates between standardized PRS and standardized PTRS in lung.

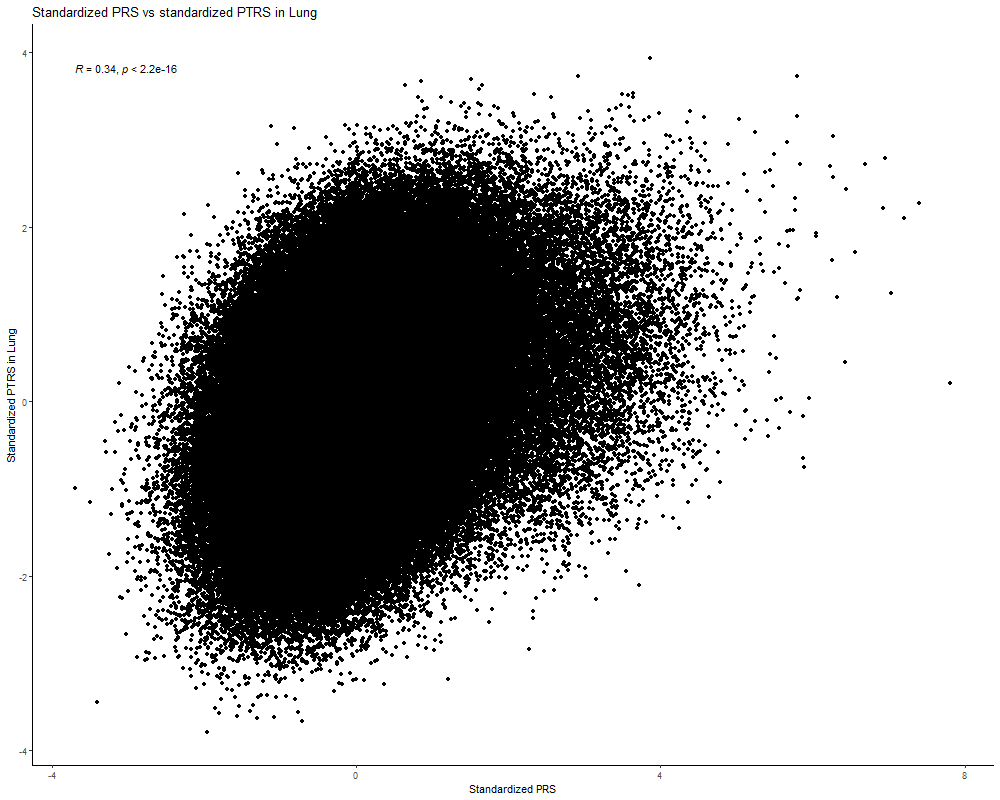

Fig. S4. Pearson correlation estimates between standardized PRS and standardized PTRS in not sun exposed skin.

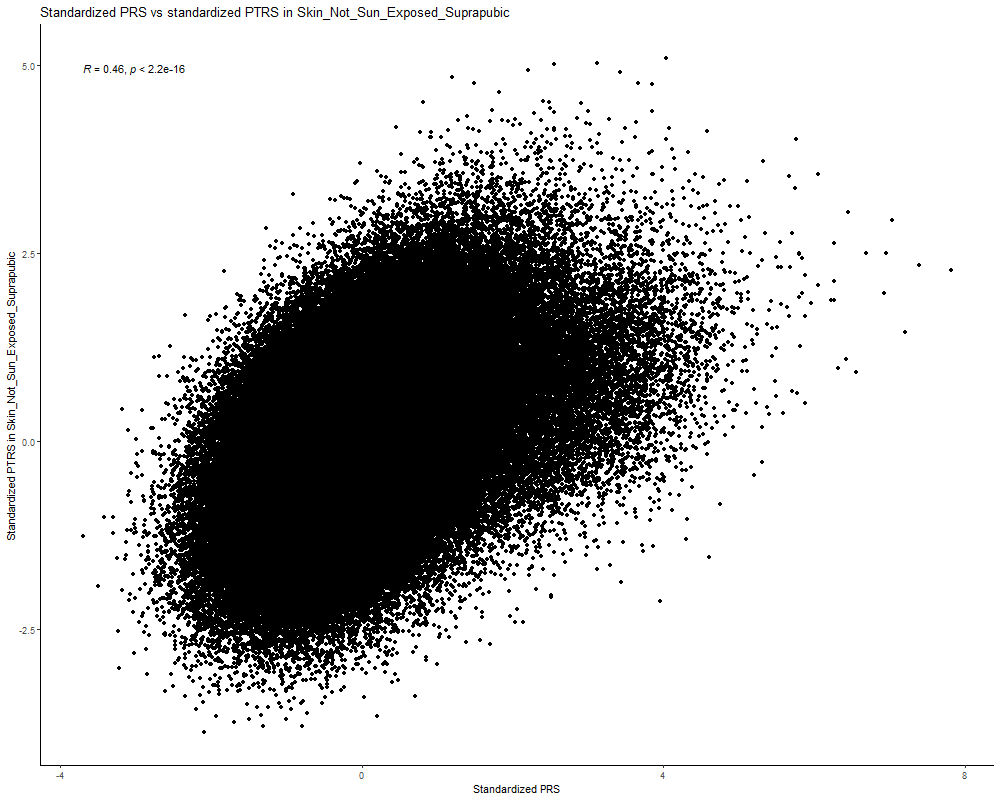

Fig. S5. Pearson correlation estimates between standardized PRS and standardized PTRS in sun exposed skin.

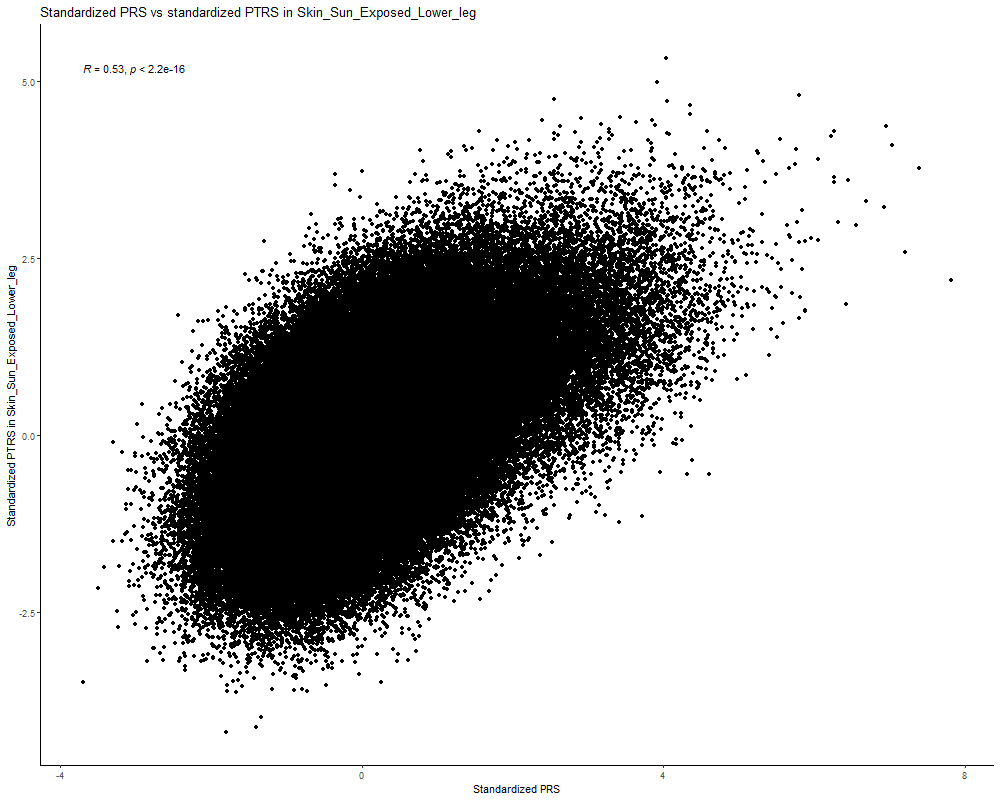

Fig. S6. Pearson correlation estimates between standardized PRS and standardized PTRS in small intestine.

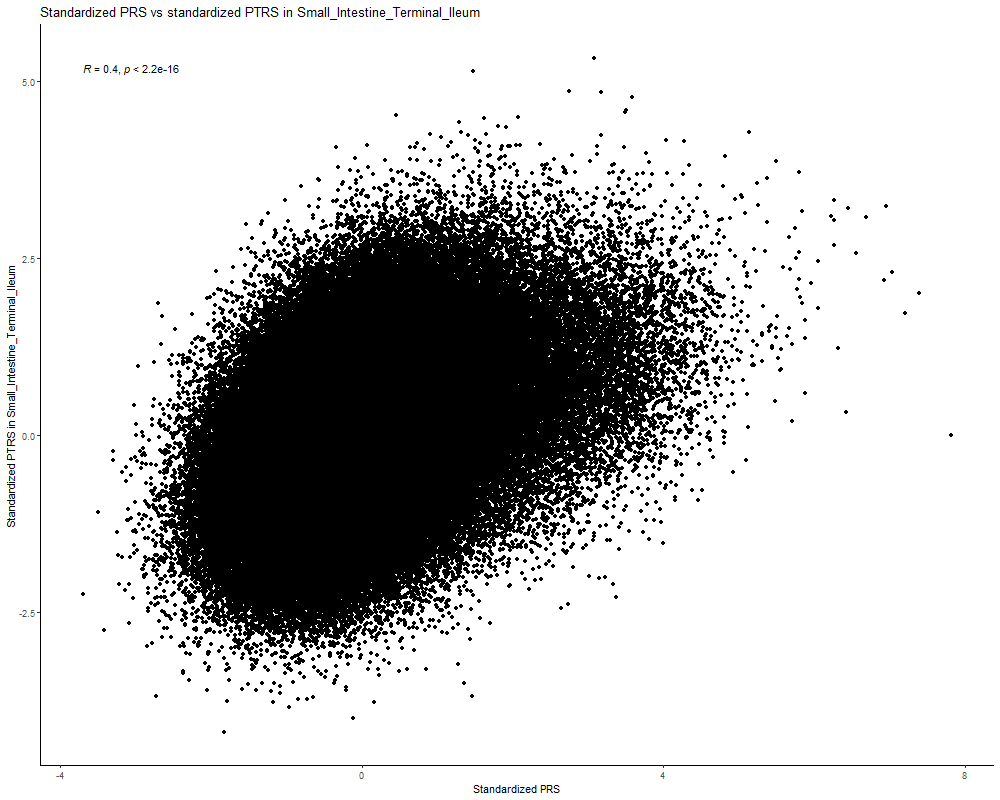

Fig. S7. Pearson correlation estimates between standardized PRS and standardized PTRS in spleen.

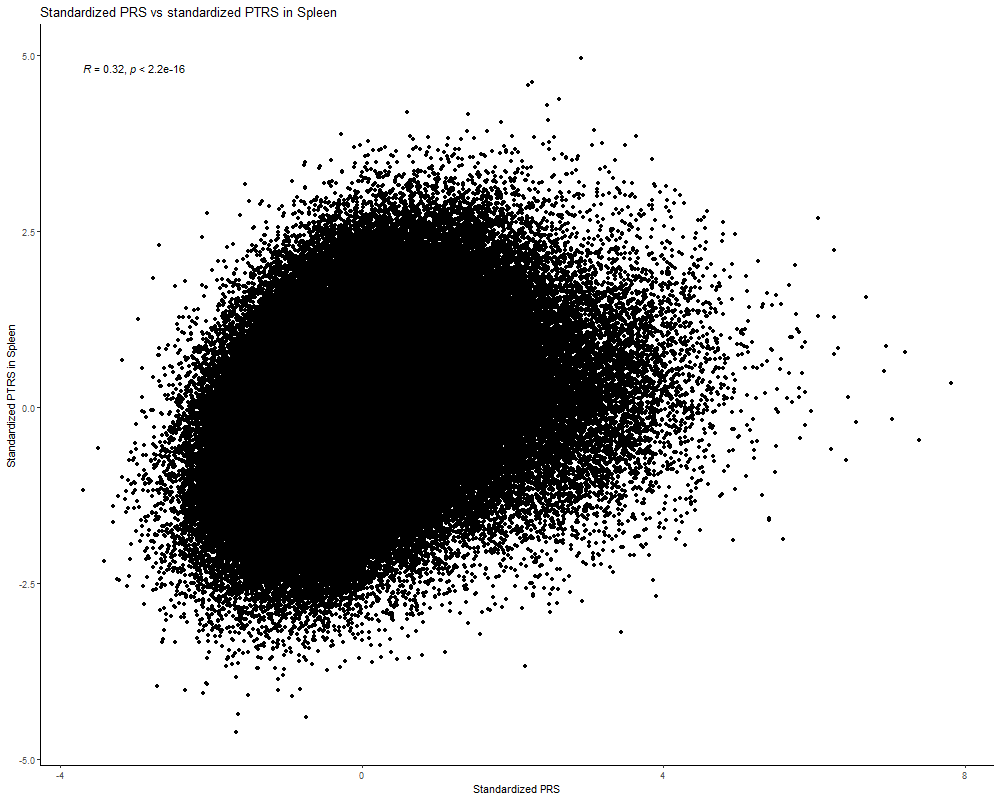

Fig. S8. Pearson correlation estimates between standardized PRS and standardized PTRS in whole blood.

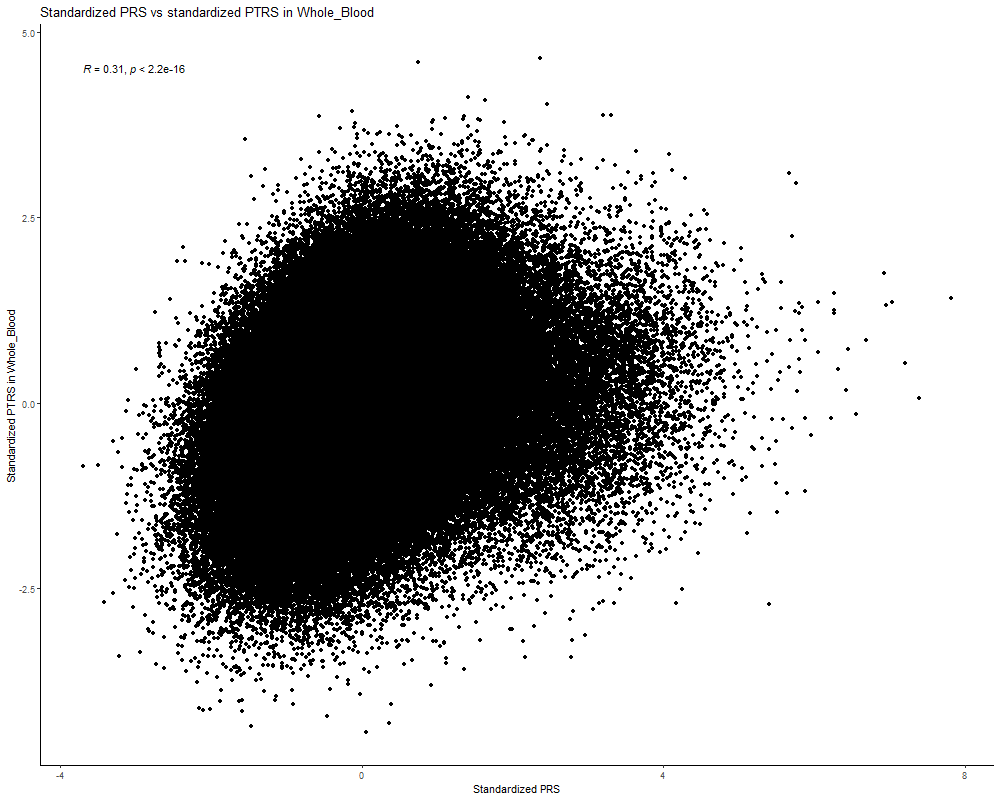
